# Model-guided placement of decentralized diagnostics for spatial control of the 2026 Bundibugyo Ebola epidemic

**DOI:** 10.64898/2026.09.29.26364338

**Authors:** Daniel J. Klein, Kyra H. Grantz, Caroline O. Buckee, Dylan B. George, Scott F. Dowell

## Abstract

Before 2026, Bundibugyo virus disease had only two recognized outbreaks. By August 2026, delayed recognition and multi-day sample transport in eastern Democratic Republic of the Congo had contributed to a large, spatially dispersed epidemic. We hypothesized that model-guided placement of decentralized diagnostics in frontier health zones could help contain spatial spread by acting as a spatial fence: delayed recognition would be converted into timely polymerase chain reaction (PCR) confirmation, isolation, and contact tracing before imported lineages became established.

We fit a Bayesian spatial branching process to public situation reports through 2 August 2026, with Flowminder movement and a latent-burden anchor. Lineages had overdispersed offspring, PCR access and turnaround, clinical recognition, isolation, tracing, safe burial, and local behaviour change. Because the observed history did not identify future control, we assumed a national response ramp yielding West-Africa-scale status-quo burden and replayed 2,000 posterior trees with 5-100 zone-level PCR laboratories, deployment delays of 0-180 days, and postmortem rapid diagnostic test (RDT) coverage.

The posterior reproduced case, death, latent-burden, and spatial targets. Speed dominated: 35 PCR laboratories averted a median 49% of future deaths if deployed immediately, 38% after 14 days, and 11% after 60 days, with 90% prediction intervals of 30-65%, 23-53%, and 5-20%, respectively. Placement also mattered: with a 30-day delay, model-informed placement averted 18-33% of future deaths with 20-100 laboratories, versus 1-8% under random placement. Postmortem RDTs partly filled gaps left by live-case detection, increasing future deaths averted from 26% to 41% with 20 laboratories, a 14-day delay, and 75% coverage. Decentralized diagnostics can slow spatial spread when they are placed ahead of likely importations and paired with staffing, isolation, tracing, and safe-burial capacity.

## 1. Introduction

Bundibugyo virus (BDBV) was first recognized as a cause of human Ebola disease after the 2007-2008 outbreak in western Uganda. The only other recognized BDBV outbreak was in 2012 in Isiro, Democratic Republic of the Congo (DRC) (MacNeil et ah, 2010; Roddy et ah, 2012; Albarino et ah, 2013; Kratz et al., 2015). Those events were small relative to the ongoing 2026 outbreak: the 2007-2008 Uganda outbreak produced 149 suspected cases and 37 deaths under one published case definition, while the 2012 DRC outbreak produced 38 laboratory-confirmed cases and 13 deaths (MacNeil et al., 2010; Towner et al., 2008; Kratz et al., 2015).

By 2 August 2026, the public INSP/INRB-UMIE record used by this project contained 3,802 confirmed cases and 1,707 confirmed deaths across 51 health zones, making the 2026 epidemic unprecedented for BDBV in both absolute burden and geographic scale.

By the time the first public analyses were released in May 2026, the combination of suspected deaths in DRC and imported cases detected in Uganda already implied substantial undetected transmission: an Imperial College London analysis estimated that roughly 400-800 infections could have occurred in DRC by 17 May 2026 (McCabe et ah, 2026). The EpiForecasts BVDOutbreakSize analysis, updated as new situation reports were released, subsequently inferred a March start and an infection burden several times larger than the laboratory-confirmed case count (Abbott et ah, 2026).

Two features of this outbreak make decentralized diagnosis particularly important. First, most early infections occurred in health zones without nearby polymerase chain reaction (PCR) laboratories. At the start of the response, samples from peripheral health zones were routed to Kinshasa or to a small number of provincial laboratories, creating turnaround times long enough for patients to reach the late, highly infectious phase of illness before confirmation. Second, spatial spread changes the value of a test. A machine in an established hot spot can still speed isolation locally, but a machine placed ahead of the frontier can prevent an imported case from growing into a local cluster of infections. This strategy is operationally plausible because cartridge-based PCR platforms and emerging rapid diagnostic tests can extend testing beyond centralized reference laboratories without requiring a full high-containment virology facility at each site. In this sense, decentralized PCR laboratories and postmortem rapid diagnostic tests (RDTs) can act as a diagnostic fence: they do not stop movement, but they can shorten the window during which a new cluster remains invisible or prevent it from taking hold in the first place. A diagnostic fence may be especially important at a mobile outbreak frontier, both for protecting newly invaded health zones and for reducing the chance of cross-border spread into vulnerable neighboring countries such as South Sudan or the Central African Republic.

That framing complements, rather than replaces, control at the epidemic core. Large hot spots still require treatment capacity, contact follow-up, community engagement, and rapid diagnosis. The diagnostic-allocation question is whether some of the limited laboratory and burial-detection capacity should also look beyond the core toward zones at risk of future importation, so the response is not always chasing new clusters after they have already established.

Mathematical models of Ebola control have repeatedly shown that no single response measure is sufficient once transmission is geographically dispersed. Models of West African and DRC outbreaks have combined isolation, contact tracing, safe burial, spatial spread, population mobility, ring vaccination, community engagement, and targeted vaccination to ask where limited response resources should be placed (Kucharski et al., 2016; Ajelli et al., 2016; Kraemer et al, 2019; Huber et al., 2023; Jombart et al., 2026). These studies provide the conceptual basis for the present analysis: Ebola response is spatial, stochastic, and delay-sensitive. The new question in 2026 is how much diagnostic infrastructure itself can change the spatial process, both by returning PCR results quickly enough to isolate living cases and by making postmortem results available immediately to families asked to consent to safe and dignified burial practices.

Quantitative policy support for this outbreak was accelerated by several changes acting together. INSP generated machine-readable situation-report data (Institut National de Sante Publique, 2026), and national mobility flows from Flowminder were available immediately. AI-assisted coding was also entering real-time outbreak modeling; the EpiForecasts BVD-OutbreakSize nowcast disclosed language-model drafting under human oversight (Abbott et ah, 2026).

Building on these broader advances, we integrated language-model coding agents throughout the present analysis. Agents were used to extract and double-check PDF tables, write and debug model code, generate figures, draft and revise manuscript text, and maintain a structured simulation experiment ledger. Human scientists set the research questions, structural assumptions, scenario definitions, and acceptance checks. Together, immediate access to data and mobility estimates, prior outbreak-modeling experience, and agent-assisted implementation reduced the time to first decision-facing results to under two days; comparable analyses had taken us weeks to months in previous outbreaks. This workflow also allowed us to develop and maintain a rapidly updated spatial branching-process model and evaluate whether diagnostic deployment should chase observed incidence or be placed ahead of the forecast transmission frontier.

We focus on three operational findings. First, speed matters: the value of local diagnosis decays within weeks, because the outbreak frontier moves and the assumed national response removes the late tail. Second, placement matters: model-informed placement at the forecast frontier reaches high-burden zones that random allocation misses. Third, rapid postmortem testing matters: it adds an independent safe-burial pathway for patients who die before live-case PCR or clinical detection.

## 2. Methods

### 2.1 Data

The model was calibrated to the full public INSP/INRB-UMIE 2026 DRC BDBV time series, pinned locally at BDBV2026-Data commit `60f6701 (build-2026-08-05-5e9266d`, SitRep 81; data to 3 August 2026). The calibration horizon was 2 August 2026, at which point the model matched against 3,802 cumulative confirmed cases and 1,707 cumulative confirmed deaths.

Spatial spread used the HDX Flowminder DRC origin-destination matrix from March 2026. That matrix covers the national health-zone system rather than only the initial outbreak provinces. The experiments analysed here used 470 health zones, comprising the Flowminder footprint, zones with observed cases, and the seed zone. Province-level event rates from ACLED (Armed Conflict Location & Event Data), accessed 5 August 2026, were used to derive the fitted operating-disruption score: events over the 12 months ending 25 July 2026 were divided by provincial population, transformed into a normalized 0-1 score, and assigned to all health zones in the province (Raleigh et ah, 2023). Laboratory activation dates, approximate PCR turnaround tiers, early Bunia and Rwampara capacity, and Ituri PCR throughput anchors reported after declaration of the Public Health Emergency of International Concern were compiled from the public situation-report laboratory sections.

The calibration also used an external latent-burden anchor from the EpiForecasts BVD-OutbreakSize analysis, which estimates infections, onsets, deaths, reporting fractions, and a time-varying reproduction number from the INSP situation reports and Uganda export data. The anchor constrains the otherwise weakly observed ratio between confirmed cases and true infections.

### 2.2 Spatial Branching Process

The epidemic was simulated as an individual-level stochastic branching process. Each infection has a parent, an infection date, a health zone, an incubation period, an illness duration, a fatal or nonfatal outcome, PCR and clinical detection states, an isolation state, a safe-burial state, and a list of its secondary infections. Susceptible depletion was ignored, which is a reasonable approximation at the fitted attack rates and health-zone populations, but a limitation for very prolonged or very large future trajectories.

The number of secondary infections generated by a case was drawn from a negative binomial distribution with mean `RO` and fixed dispersion `k`, allowing superspreading. Offspring infection times were assigned from a gamma generation-interval distribution with mean 15.3 days and standard deviation 9.3 days, matching the West African Ebola Zaire serial-interval estimate used as a generation-time proxy by WHO Ebola Response Team and the BVD-OutbreakSize model (WHO Ebola Response Team, 2014; Abbott et al., 2026). Incubation periods were lognormal with mean 6.3 days and standard deviation 3.5 days, using the BDBV incubation mean from the 2007-2008 Uganda outbreak (MacNeil et al., 2010). Fatal infections had onset-to-death times drawn from a gamma distribution with mean 12 days; nonfatal infections had onset-to-recovery times drawn from a gamma distribution with mean 16 days. These illness-duration parameters, as well as the assumption that 37% of a fatal case’s transmission potential can fall in a 0.8-day postmortem period, were fixed naturalhistory assumptions informed by Ebola case-series data and field interpretation and varied only in scenario sensitivity checks; fixed values, priors, and posterior summaries are listed in Appendix.

Infectiousness was concentrated late in disease. For each case, daily infectiousness rose linearly from symptom onset to the illness endpoint. For fatal cases, an additional postmortem window represented transmission during unsafe burial. Isolation truncates the remaining live transmission after the isolation date; safe burial truncates the postmortem component.

For each offspring infection, the destination health zone was local with probability `1 -jump_prob;` otherwise the destination was drawn from the parent’s row-normalised Flowminder outflow row. The fitted `jump_prob` was shared across health zones; Flowminder flows determined the relative destination probabilities conditional on a between-zone jump. Zonespecific transmission multipliers derived from a province-level operating-disruption score allowed realised growth to vary across the country.

**Figure 1:**
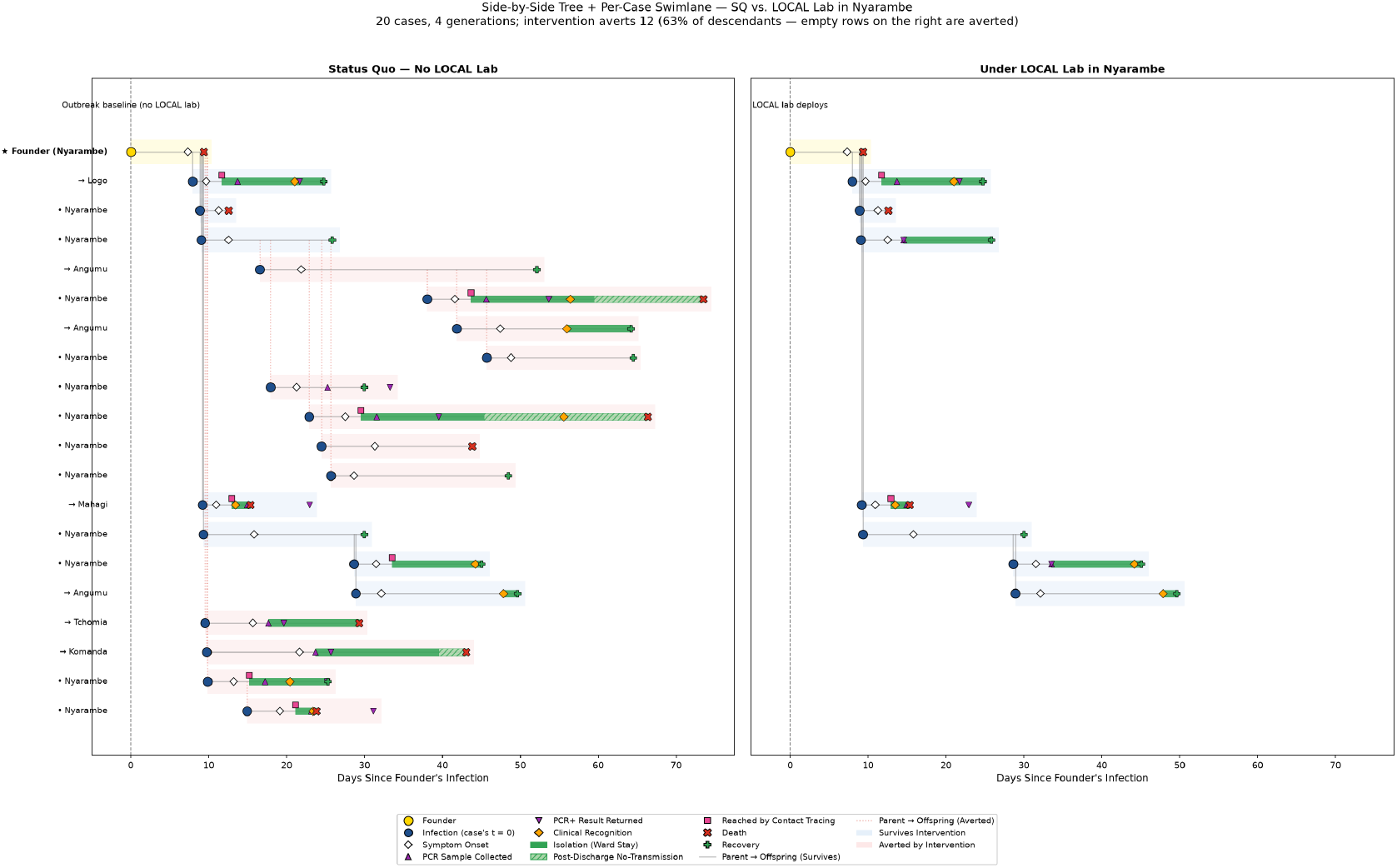
Spatial branching process and diagnosis timeline. Infections are explicit nodes in a transmission tree. PCR confirmation, clinical recognition, isolation, contact tracing, and safe burial cut different parts of a case’s potential offspring. In this illustrative branch, a local laboratory in Nyarambe leads to detection and isolation before a jump from Nyarambe to Angumu that would have generated eight infections; safe and dignified burial then averts three direct infections and one onward infection that would have occurred after death. The local laboratory does not eliminate all forward transmission, but it substantially reduces spread even over the four generations shown.

### 2.3 Behavioral And National Response

Local behavior change was represented with a Hill function of lagged visible deaths in each zone. The per-zone brake reduced transmission toward a fitted local floor as cumulative local mortality accumulated:

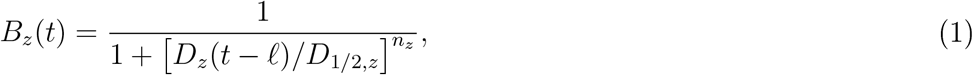

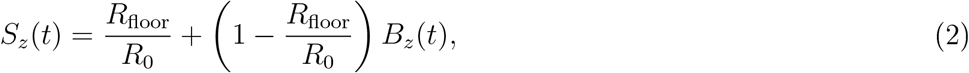

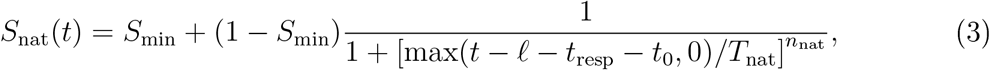

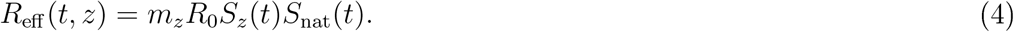

Here *D*_*z*_ is cumulative post-recognition infections in health zone *z*, multiplied by the fitted infection fatality ratio and lagged by *l* =16 days so that the driver approximates visible deaths; *D*_*1*_*/*_2_,_*z*_ and *n*_z_ are the fitted local half-saturation and fixed Hill-shape parameters; *S*_z_ maps the Hill brake onto the fitted interval between *R*_0_ and *R*_floor_; and *m*_*z*_ is a zone multiplier derived from the province-level operating-disruption score. We introduced a possible national reduction in transmission after the 2 August 2026 calibration horizon to represent broad behavioral change and non-diagnostic response activities that would be invisible to the likelihood. The parameters governing this ramp-down were chosen against a 540-day forward anchor based on the 2014-2016 West Africa Ebola Zaire confirmed-case burden. This choice produced one plausible future at roughly West-Africa scale for the dcccntralizcd-diagnostic analysis; it was not a posterior estimate of future response strength. The primary scenario reached half of its effect after 90 days and used a final scale of 0.25 with Hill exponent 2. All diagnostic effects below arc conditional on that ramp and on the PCR and RDT assumptions in each scenario.

**Figure 2:**
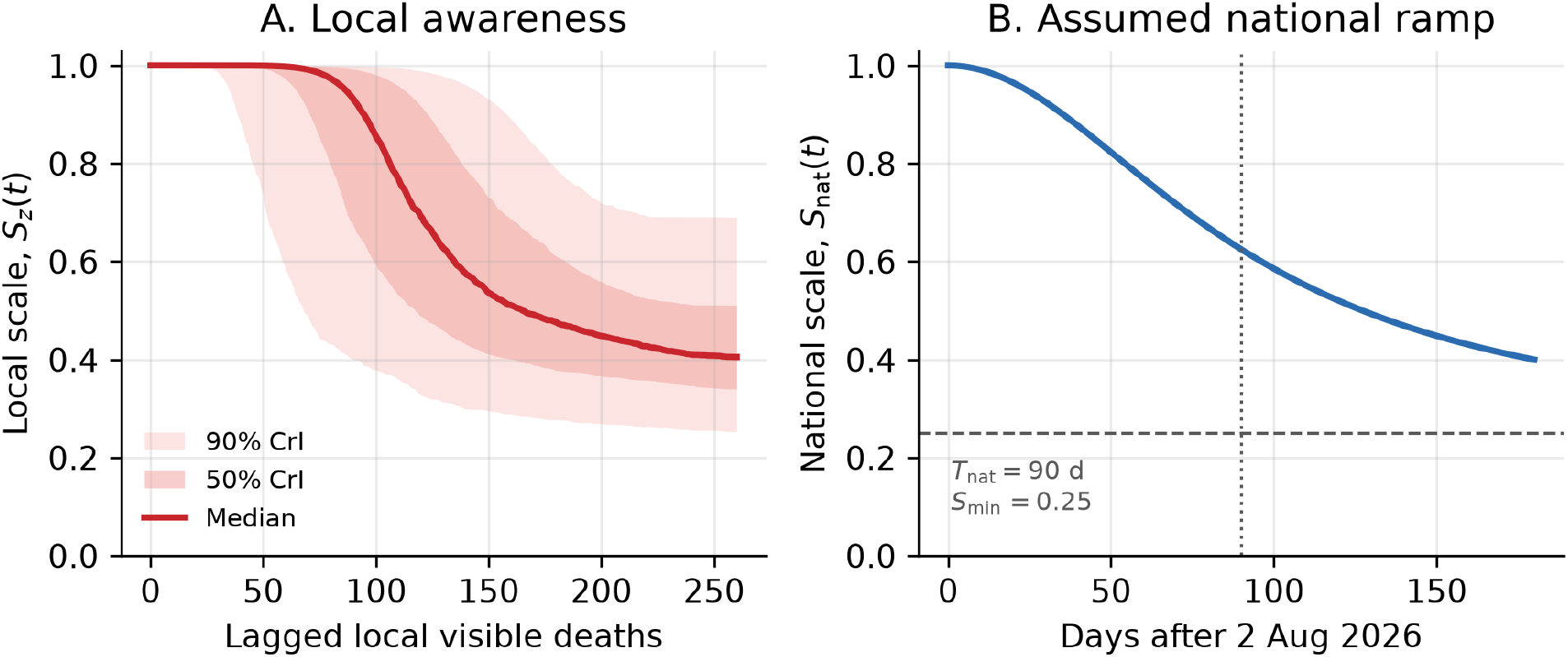
Local and national response functions. The local scale, *Sz*(*t*), fell as cumulative lagged visible deaths accrued in each health zone; shading shows 5-95% and 25-75% posterior intervals propagated through *R*_0_, *R*_floor_, and *D*_1/2,z_, with the Hill shape fixed at 8.7. The postcutoff national scale, *S*_nat_(*t*), was fixed to the selected *T*_nat_ = 90 d, *S*_min_ = 0.25, *n*_nat_ = 2 scenario rather than fitted.

### 2.4. Diagnosis, Isolation, Tracing, And Safe Burial

Living cases could be detected through PCR testing and through a separate clinical recognition channel. PCR access was represented as a three-tier ladder: high access for served health zones with a capacitated laboratory, intermediate access for aware non-lab zones in which prior local confirmations had raised suspicion, and low access for BDBV-naive unserved zones without local PCR access or recent confirmed cases. The primary analysis assumed that routine sample transport between health zones was highly challenging: a PCR laboratory primarily served the health zone in which it was placed. Health zones with status-quo laboratories were therefore treated as already served and were excluded from new laboratory placement, leaving 462 candidate zones.

PCR sample collection occurred after onset, with shorter symptom-to-sample delays in served or aware zones. Samples from unserved zones incurred a long turnaround time for shipping to a centralized lab in Kinshasa; samples from lab zones returned on a one-day local turnaround. A PCR-positive living case could be effectively isolated first as a suspect and then, if not already isolated, as a confirmed case. The confirmed isolation probability was parameterized as an ordered uplift over presumptive isolation:

~~~
p_iso_confirmed - p_iso_presump + (1 - p_iso_presump) * iso_uplift
~~~

so confirmation cannot reduce the probability of isolation. PCR sensitivity was not modeled as a separate failure process: a sampled true BDBV infection was assumed to be PCR-confirmed.

Confirmed cases were also eligible to trigger contact tracing. Contacts who were the direct offspring of a confirmed index case were traced with probability `sq_trace_eff` and isolated after a two-day tracing lag. This representation treats tracing as an offspring-level probability rather than as an explicit contact graph.

Fatal cases that had been diagnosed during life were safely buried. Fatal cases who died undiagnosed could be detected by a postmortem rapid diagnostic test. The status quo retained each posterior draw’s baseline postmortem PCR coverage. RDT scenarios then increased postmortem testing to at least 50% or 75% coverage of otherwise undiagnosed deaths and assumed 80% sensitivity. Decedents with a positive RDT were assumed to receive a safe and dignified burial without additional modeled delay, so the channel remained imperfect but a positive test was actionable. This use case is intentionally peri-mortem: Ebola RDT sensitivity is highest when viral loads are extremely high late in illness or after death, so the RDT intervention complements, rather than substitutes for, PCR confirmation earlier in the disease course.

### 2.5 Calibration

The status quo was calibrated with a pseudo-marginal MCMC sampler (Andrieu and Roberts, 2009) implemented with `emcee` (Foreman-Mackey et ah, 2013). The sampler included 14 parameters and a latent random seed for each simulated tree. Because the likelihood is stochastic, each parameter vector was evaluated by averaging over 10 independent simulator seeds; a single tree was then selected within each parameter state with probability proportional to its likelihood contribution. This yielded samples from the joint posterior over parameters and realised epidemic trajectories, which could be replayed exactly under matched counterfactuals.

The sample-transport refit was targeted for 60,000 steps per arm; the intervention sweep reported here used a thinned checkpoint from iteration 44,000 of the health-zone-only arm. We extracted 2,500 posterior rows, randomly reserved 500 for the greedy placement pre-pass, and continued 2,000 disjoint held-out posterior states five times under the status quo and under each diagnostic scenario to estimate intervention effects.

### 2.6. Counterfactual Diagnostic Scenarios

*\Nc* evaluated newly capacitated PCR laboratories placed in health zones not served by status-quo laboratories. Budgets were 5, 10, 20, 35, 50, 75, or 100 laboratories. The modeled unit was a deployable cartridge-based PCR node, such as the GeneXpert-or RADIOXE-class platforms used during the 2026 response, rather than a full central reference laboratory. We considered two mechanisms by which new local molecular capacity affected access to diagnosis. In the primary local-placement mode, a laboratory improved diagnosis in the health zone where it was placed, with three modeled effects: higher PCR sample-collection probability, earlier sample collection after symptom onset, and one-day local turnaround rather than the long centralized shipping delay for unserved zones. In a sample-transport sensitivity analysis, unserved health zones could ship specimens to the nearest new laboratory, receiving an intermediate PCR access probability and a road-time-based turnaround delay rather than full local-lab access. A lazy CELF optimizer (Leskovec et ah, 2007) selected laboratories by marginal deaths averted under common random numbers; random placement was used as the comparator.

Each diagnostic scenario was evaluated by replaying the same posterior tree used for the matched status-quo simulation. Both simulations shared the same per-case random numbers; the intervention pruned only transmissions that would have been prevented by an earlier PCR result, isolation event, traced contact, or safe burial.

Deployment delays of 0, 14, 30, 60, 90, 120, and 180 days after the calibration horizon measured the penalty for late roll-out. Postmortem RDT scenarios raised baseline postmortem testing to 50% or 75% coverage of otherwise undiagnosed deaths, either inside the PCR-machine footprint or everywhere. Unless otherwise noted, reported percentages use the **post-cutoff still-avertable** denominator: deaths that would occur after 2 August 2026 under the simulated status quo.

### 2.7 AI-Assisted Implementation And Audit

The analysis was implemented with human-in-the-loop AI support. Claude Opus was used for situation-report translation and extraction, Claude Code Opus for simulation engineering and figure generation, and OpenAI ChatGPT for independent review of modeling assumptions, results, and manuscript language. Human scientists set the model structure, calibration targets, scenario definitions, countcrfactual algorithms, and acceptance gates; authors reviewed and revised AI-assistcd code, figures, and text. Generated code and outputs were then constrained by reproducibility checks, including local data-pin assertions, byte-for-byte replay of matched stochastic trees, guards against pro-cutoff changes when future-only assumptions were varied, and metadata blocks that recorded denominators and posterior vintages alongside figure summaries.

## 3. Results

### 3.1 Historical Fit And Scenario Baseline

The posterior used for the intervention sweep reproduced the national confirmcd-casc curve, cumulative confirmed deaths, isolation occupancy, external latent-burden anchors, and the observed pattern of spatial spread through the calibration horizon. Its median `RO` was 2.63. median fitted infection fatality ratio was 0.46, and the realised latest `Rt` in a 200-draw diagnostic was 1.11. The median bctwccn-zonc jump probability was 0.19, implying substantial movement among small health-zone units. The median probability that a case in a lab health zone was PCR-tcstcd was 0.60, while the corresponding probability for a naive unserved zone was 0.29. Contact tracing from a PCR-confirmcd index was inferred to intercept a median 69% of its offspring.

**Figure 3:**
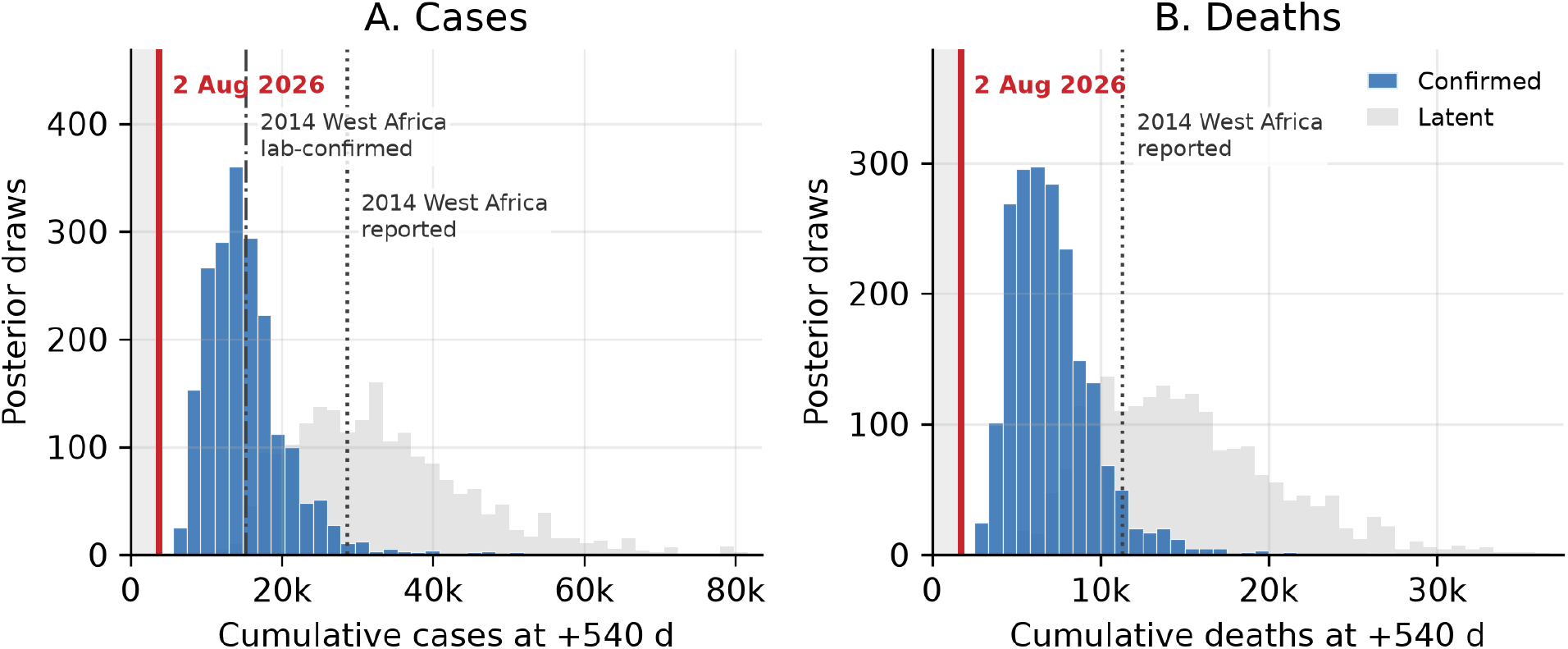
Assumed status-quo futures under the primary national response ramp. Posterior epidemic trajectories were continued for 540 days after 2 August 2026 with the selected non-diagnostic response ramp and no added diagnostic capacity. Blue histograms show confirmed cases or deaths expected to be observed by the model, gray histograms show latent infections or deaths, and the shaded region marks burden already accrued by the model horizon and therefore unavailable to future interventions. The model-confirmed case distribution is comparable with 2014 West Africa laboratory-confirmed cases; latent infections and deaths expose the much larger true epidemic burden that remains partially observed.

Under the status quo plus the selected national response ramp, the simulated outbreak reached a median of 31,615 cumulative infections, 14,256 deaths, and 110 invaded health zones by 540 days after 2 August 2026. Of those deaths, a median 4,816 had already been locked in by the calibration horizon, leaving a median 9,360 future deaths that could still be averted.

### 3.2 Speed Matters

The value of new laboratories fell steeply with deployment delay. For 35 locally placed laboratories, median deaths averted were 4,331 with immediate deployment, 3,362 with a 14-day delay, 2,370 with a 30-day delay, 1,004 with a 60-day delay, 314 with a 90-day delay, 78 with a 120-day delay, and 2 with a 180-day delay. Expressed as a fraction of deaths still avertable after the cutoff, the same curve fell from 48.6% at immediate deployment to 37.6% at 14 days, 26.6%> at 30 days, 11.1% at 60 days, and less than *1%* at 120 days.

The median delay at which the benefit of local PCR fell to half of its day-zero value was 34.5 days. This short clock is a joint property of the epidemic and the assumed national support ramp: the outbreak continued to seed new health zones for long enough that decentralized diagnostics had substantial value, but the same national ramp also removed most of the late tail by 120-180 days.

**Figure 4:**
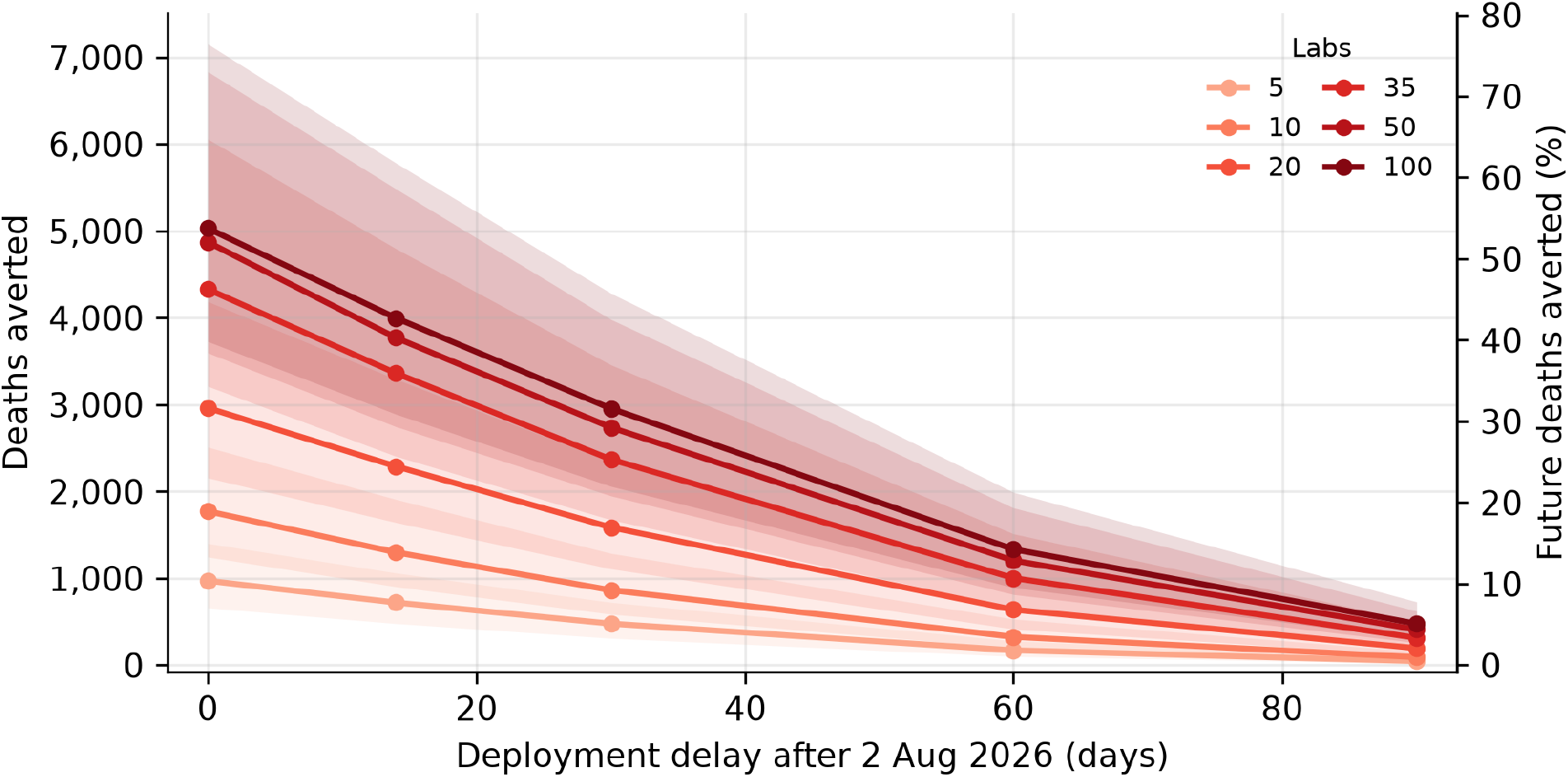
Speed dominates the deployment problem. For a fixed laboratory budget, deaths averted decline rapidly as deployment is delayed after the 2 August 2026 model horizon. Points show medians and shaded intervals show interquartile ranges across posterior draws; the right axis expresses the same absolute death scale relative to the median 9,360 future deaths remaining under the status quo.

### 3.3. Placement Matters

Forward placement dominated random allocation. With a 30-day deployment delay. 20 model-placed laboratories averted a median 18.0% of future deaths, compared with 1.2%) under random placement; 100 model-placed laboratories averted 32.3%, compared with 8.0%) under random placement. The optimized strategy beat random placement in 100%) of the 2,000 paired posterior draws at every budget tested.

The optimized placement list was concentrated on likely frontier exporters rather than only on current incidence. In the day-30 placement sweep used here, the top 15 selected zones were Beni, Logo, Kyondo, Nyarambe, Biringi, Katwa, Vuhovi, Aungba, Masereka, Mandima, Karisimbi, Mahagi, Angumu, Rimba, and Butembo.

The same day-30 plans also reduced spatial spread. In a matched 2,000-draw replay of the canonical continuation from each posterior state, the status quo seeded a median 32 new health zones after 2 August 2026. Model-placed PCR laboratories reduced that to 29 new zones with 20 laboratories, 26 with 50 laboratories, and 25 with 100 laboratories.

**Figure 5:**
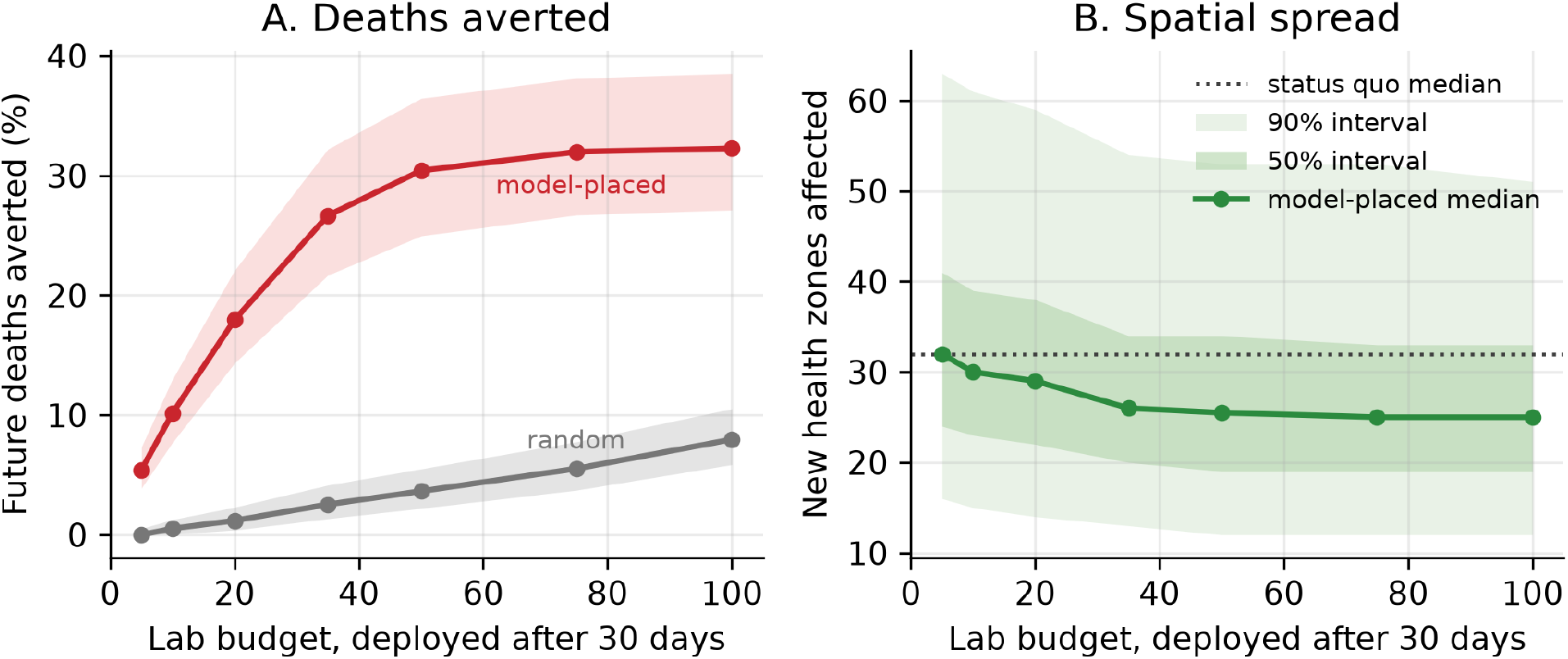
Placement matters when capacity can be scaled ahead of the frontier. Model-placed laboratories averted more deaths than random placement at the same 30-day deployment delay and reduced the median number of new health zones affected after the model horizon. Panel A summarises the full 2,000-draw day-30 placement sweep; Panel B replays the same 2,000 posterior states once each to count health zones affected after 2 August 2026 under the status quo and matched local-PCR counterfactuals, with shaded 50%) and 90%) intervals.

**Figure 6:**
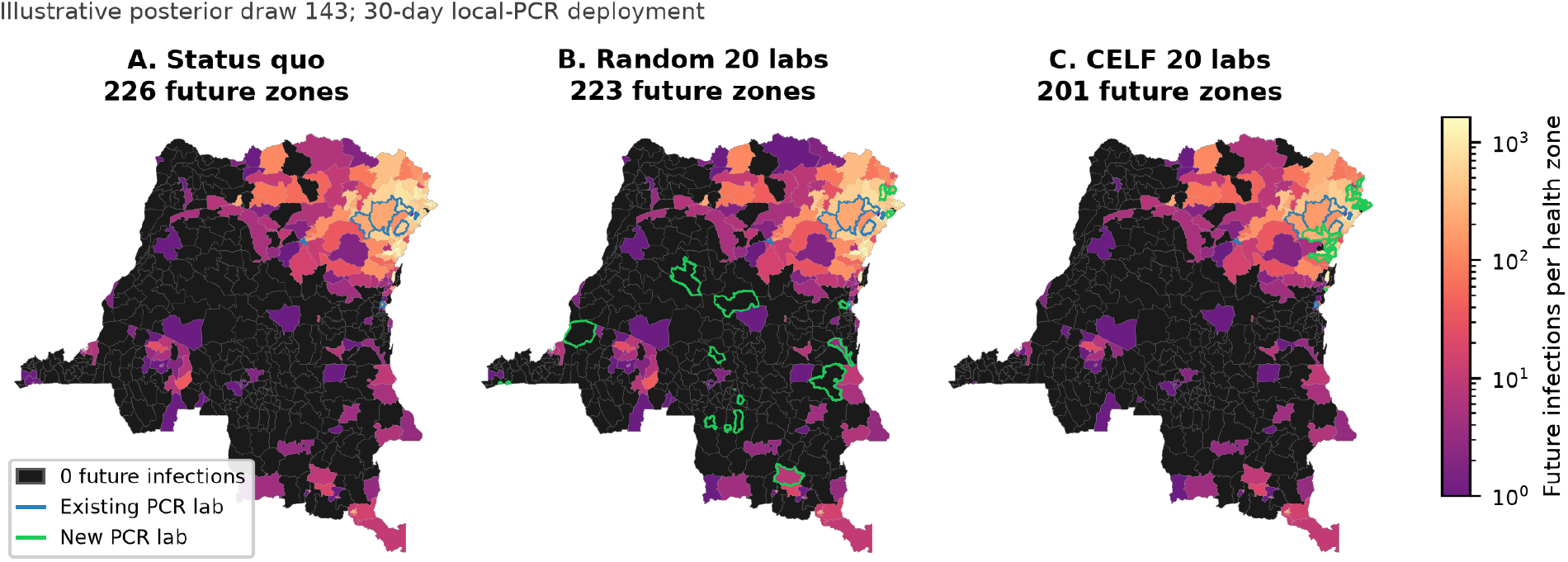
Optimized placement reduces spatial spread in one illustrative iteration of the branching process. Maps show future infections after 2 August 2026 in one posterior continuation under the status quo, a fixed 20-laboratory random allocation, and the first 20 health zones selected by the CELF-optimized local-PCR placement order. Blue outlines mark existing PCR laboratory health zones and green outlines mark newly placed laboratories in each intervention panel. The same underlying transmission tree is used in all three panels; only the transmissions pruned by the diagnostic counterfactual differ.

### 3.4. Postmortem RDT Matters

Postmortem RDTs added a safe-burial pathway that PCR placement alone could not reach. At 20 new laboratories with a 14-day delay, local PCR alone averted a median 2,290 deaths, or 25.5% of post-cutoff preventable deaths. Adding RDTs for 50% of otherwise undiagnosed deaths increased the median effect to 3,186 deaths and 34.7%; adding 75% RDT coverage increased it to 3,767 deaths and 40.7%.

The timing curves show the same pattern. At day zero, 20 PCR laboratories alone averted a median 33.5% of future deaths, 50% RDT coverage increased that to 44.3%, and 75% RDT coverage increased it to 51.4%. By 60 days, all three effects were sharply reduced, but 75% RDT coverage still nearly doubled the median fraction of future deaths averted relative to PCR alone, from 7.2% to 12.8%.

**Figure 7:**
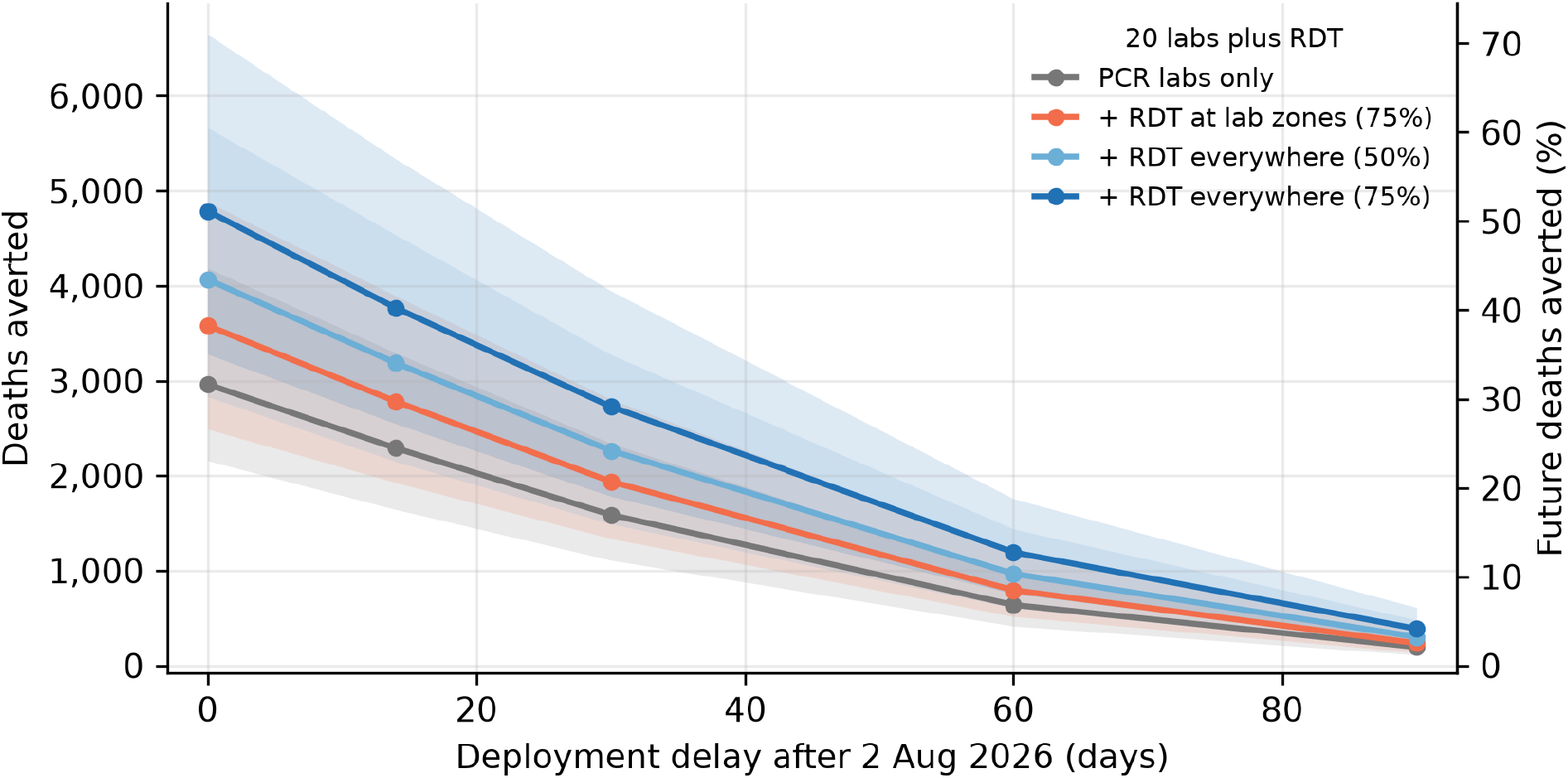
Rapid diagnostic tests add benefit through safe burial. For 20 PCR laboratories, cadaver RDT scenarios added postmortem detection for patients who were not confirmed alive, with incremental benefit over local PCR alone. Points show medians and shaded intervals show interquartile ranges across posterior draws; the right axis expresses the same absolute death scale relative to the median 9,360 future deaths remaining under the status quo.

## 4. Discussion

### 4.1. Principal Findings

These analyses support three linked conclusions. First, speed matters. A laboratory deployed after the frontier has passed still improves care and confirmation, but it no longer prevents establishment in that zone; in the primary scenario, the marginal benefit of a 35-lab package was cut by half after approximately 35 days and was almost exhausted by 120 days. The exact timing of that decline is tied to the assumed future response trajectory, but the mechanism is general. Second, allocation cannot be based only on current confirmed cases. Because the major value comes from stopping future lineages, the target is a probability distribution over future importations. Third, postmortem RDT is complementary to PCR. It reaches the fatal, undiagnosed cases that generate funeral transmission precisely because they did not pass through the live-case confirmation path.

The “diagnostic fence” analogy is useful but imperfect. Laboratories and RDTs do not form a literal cordon, and they cannot block movement. They instead change the life history of lineages that land in covered zones: PCR increases the chance and speed of live confirmation, confirmation improves isolation and contact tracing, and postmortem RDT moves an otherwise unsafe burial into the safe-burial pathway. The fence is therefore probabilistic, delayed, and leaky. Its holes are not only device failures; they are missed patients, late care seeking, absent staff, backlogged samples, untraced contacts, unsafe burials, and zones where local insecurity prevents any of the above from operating. The scenarios also assume demand for decentralized care. Many deaths were undiagnosed in the community while testing remained low; placing diagnostics and care closer to patients can only find those cases earlier if people seek care for nonspecific early symptoms. Limited care seeking would weaken this mechanism, although demand may rise if better local service makes early presentation more useful and trusted.

### 4.2. Why Al Changed The Analysis Timeline

AI changed the analysis timeline by compressing the tedious middle of the workflow rather than by replacing scientific judgement. Language-model agents helped translate and reconcile French PDF SitReps, implement paired replay tests, audit posterior and denominator metadata, draft and revise manuscript text, and revise figures as assumptions changed. The same process exposed several mistakes: scenario scripts that inherited a national ramp from an older posterior, a deaths figure initially rendered from an infection-denominator summary, and profile experiments that failed to rescue assumptions we expected to work. The durable workflow was therefore not autonomous forecasting, but rapid Al-assisted implementation bounded by human model design, data hashes, common-random-number replay, cross-machine likelihood checks, and review of the resulting epidemiology.

### 4.3. Relation To Previous Ebola Models

Previous Ebola models established that stochastic extinction, superspreading, contact tracing, burial safety, and geography are all central to control. The matched replay design used here applies that branching-process perspective to diagnostic counterfactuals, turning faster PCR, isolation, contact tracing, and safe burial into explicit cuts in a shared transmission tree.

The mobility component plays a role analogous to metapopulation models used for preparedness and point-of-control placement, but at the health-zone level and with CDR-derived Flowminder movement probabilities. That movement layer is what turns diagnosis from a local clinical intervention into a spatial allocation problem. Immediate access to health-zone mobility estimates also made it possible to estimate where the next outbreak frontiers could emerge rather than only react to observed incidence, underscoring the preparedness value of similar mobility layers in at-risk neighboring countries such as South Sudan and the Central African Republic. It also creates the main per-zone interpretability caveat: the model places cases where infection occurred and does not include explicit referral to regional hospitals, nosocomial amplification, or facility-level transmission.

### 4.4. Operational Implications

The response required by the model is broader than placing boxes on a map. Decentralized PCR capacity is useful only if samples can be collected in the first few days after onset, results can be returned fast enough to change behavior, isolation units can take patients, and confirmed cases trigger contact follow-up within days. Postmortem RDTs create an analogous benefit only if a positive result immediately changes burial practice. The SitRep record used here showed persistent stress on contact tracing and isolation occupancy, including contact follow-up rates below stated operational targets and many confirmed cases outside known contact lists. Those constraints make laboratory placement necessary but not sufficient.

The sample-transport analysis also changes the practical interpretation of where laboratories matter. If a zone assigned to an existing laboratory is not served quickly in practice, the model will incorrectly assign it a high PCR probability and then conclude that adding a laboratory there has little value. Real catchments should be measured as an operational object: cold-chain time, road quality, fuel, sample pick-up frequency, security, staffing, and result-return latency, not map distance alone.

The placement result should also be interpreted as a fixed allocation problem. The optimizer was given the posterior at one decision time and asked where a fixed number of laboratories should go. Field placement is more reactive: response teams learn where new clusters are occurring, which roads are passable, and which laboratories have become backlogged before the next tranche of capacity arrives. Future analyses should therefore evaluate reactive placement policies that assimilate the latest epidemiologic intelligence rather than only ranking a single up-front batch.

The placement objective was also national: it minimized expected burden inside DRC. An international objective could deliberately put additional weight on northern and northeastern frontier zones if preventing cross-border exportation into the Central African Republic or South Sudan were to be prioritized. That modification would require mobility layers and response assumptions for neighboring health systems, but it is conceptually straightforward: the value of a diagnostic fence depends on what lies behind it, not only on where the next infections are most numerous.

Finally, RDT should not be framed as an inferior substitute for PCR. In this model it answers a different question: what happens after an undiagnosed person dies or is close to death? RDTs have sufficient sensitivity only near the end of disease, when viral loads are very high, but that is exactly where PCR has often already failed to change the outcome for an undiagnosed fatal case. In a disease where infectiousness rises toward death and burial can generate secondary cases, point-of-burial detection can remove transmission that no live-case laboratory ever saw. The RDT result still needs a trained burial team and community acceptance to become transmission control.

### 4-5. Limitations

The model is intentionally stylized. It ignores susceptible depletion, omits onward transmission after importations to neighboring countries, represents contact tracing as a probability on true offspring rather than an explicit observed contact network, has no healthcare-facility compartment or nosocomial transmission channel, and routes new infections through Flow - minder movement rather than through patient referral. Those choices keep the simulation fast enough for posterior counterfactuals but soften facility-level siting conclusions. The model should be interpreted as ranking broad frontier risk and mechanism sizes, not as a deterministic list of which exact clinics must receive the next devices.

The status quo also contains a national response ramp chosen against a 540-day West Africa confirmed-case anchor. That clock represents non-diagnostic support arriving after the calibration horizon: staff, safe-burial teams, treatment capacity, coordination, community engagement, and broad behavioral change. It is chosen, not fitted, because the likelihood is exactly blind to its parameters inside the fitted window. All diagnostic effects reported here are therefore marginal to that assumed national ramp.

The size of the baseline future also affects the numerical scale of every counterfactual. If nondiagnostic control were much faster than assumed here, the epidemic would be smaller and fewer deaths would remain for decentralized diagnostics to avert. If transmission were larger or more sustained, the window for PCR placement would widen and spatial diagnostics could have more absolute impact. We expect the relative ranking of strategies to be less sensitive than the absolute magnitude of benefit: speed, placement, and postmortem gap-filling all follow from diagnosis breaking future transmission chains, but the delay curves and optimal laboratory budgets would change across smaller and larger futures.

Postmortem RDT is modeled as coverage multiplied by sensitivity and affects only safe burial. We do not represent faster notification from postmortem positives back to the local awareness process because the counterfactual replay engine does not propagate posthoc state changes across generations except through offspring pruning. This makes RDT effects one-sided lower bounds for any pathway that would also accelerate community recognition or contact tracing.

The intervention analysis was also limited to one primary sample-transport assumption. Rerunning the scenario package across intermediate catchment brackets would show how sensitive the placement ranking is to partially functional inter-zone specimen transport, result return, and laboratory catchment operations.

Finally, the model considered epidemiologic benefit but not cost. It can estimate marginal infections, deaths, and health-zone invasions averted by decentralized PCR or postmortem RDT, but it cannot determine whether a particular mix of laboratories, staff, sample transport, and burial teams is cost-effective.

## 5. Conclusion

By August 2026 the DRC Bundibugyo outbreak was already the largest recognized BDBV outbreak and among the largest Ebola outbreaks ever recorded, exceeded in confirmed cases only by the 2014-2016 West Africa epidemic. It was still spreading spatially and unfolding without a licensed BDBV-specific vaccine or therapeutic. The core result of this analysis is that decentralized diagnostics can have substantial value when they are model-guided and placed on the forecast frontier, not only where confirmed incidence is already high. Forward placement reduced future deaths and new health-zone invasions in the scenarios tested here, consistent with the mechanism: once an invisible lineage crosses the frontier, timely local confirmation can prevent it from becoming a new source.

The model also argues for pairing PCR and RDT. PCR reduces live transmission and creates the confirmed indices needed for contact tracing; postmortem RDT covers the deaths that were missed during life and converts unsafe burials to safe burials. But neither intervention is autonomous. The scale of diagnostic roll-out has to be matched by the human system that makes a result actionable: sample transport, laboratory staff, result return, isolation capacity, contact tracing, and safe-burial teams.

Because the outbreak was still growing at the model horizon, next steps should move from fixed placement to adaptive roll-out, add the costs of the laboratories and field teams needed to make diagnosis actionable, and prepare the same modeling machinery for neighboring countries if the epidemic seeds new cross-border transmission.

## Data Availability

This paper contains simulation-based disease modeling of the BDBV outbreak. Source code is available online.

https://github.com/InstituteforDiseaseModeling/bundibugyo-decentralized-diagnostics

## 6. Code Availability

Source code for the minimal reproduction package accompanying this preprint is available in the `bundibugyo-decentralized-diagnostics` repository under the Institute for Disease Modeling GitHub organization.

## Appendix 1: Key Parameters

Table Al lists the natural-history assumptions fixed in the model and selected calibrated parameters from the posterior used in the diagnostic scenario sweep. Posterior summaries are medians with 5–95% intervals from the health-zone-only posterior propagated into the diagnostic scenario sweep.

**Table 1.**
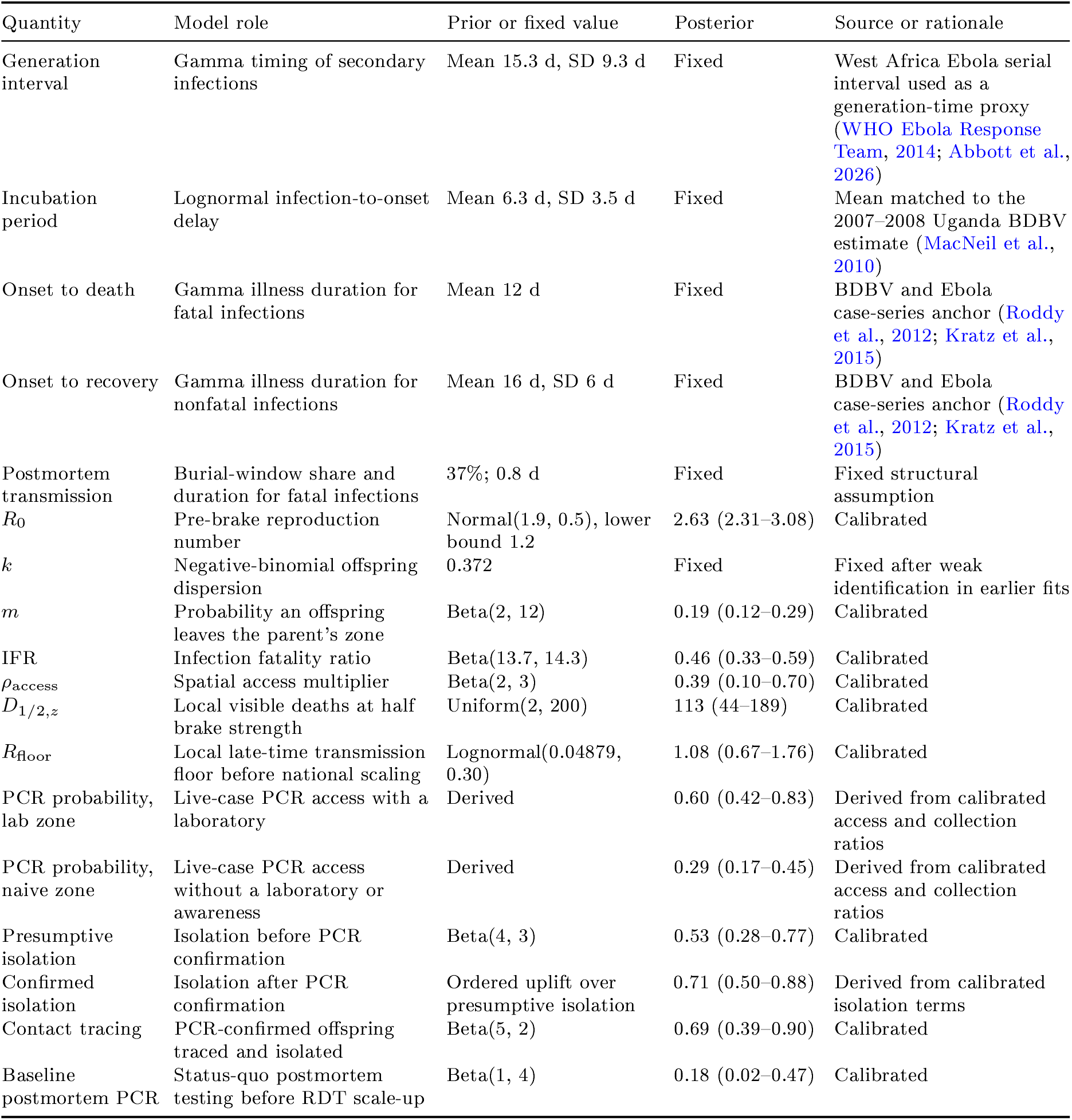
Key fixed assumptions, priors, and posterior summaries used in the diagnostic scenario analysis.

| Quantity | Model role | Prior or fixed value | Posterior | Source or rationale |
| --- | --- | --- | --- | --- |
| Generation interval | Gamma timing of secondary infections | Mean 15.3 d, SD 9.3 d | Fixed | West Africa Ebola serial interval used as a generation-time proxy (WHO Ebola Response Team, 2014; Abbott et al., 2026) |
| Incubation period | Lognormal infection-to-onset delay | Mean 6.3 d, SD 3.5 d | Fixed | Mean matched to the 2007–2008 Uganda BDBV estimate (MacNeil et al., 2010) |
| Onset to death | Gamma illness duration for fatal infections | Mean 12 d | Fixed | BDBV and Ebola case-series anchor (Roddy et al., 2012; Kratz et al., 2015) |
| Onset to recovery | Gamma illness duration for nonfatal infections | Mean 16 d, SD 6 d | Fixed | BDBV and Ebola case-series anchor (Roddy et al., 2012; Kratz et al., 2015) |
| Postmortem transmission | Burial-window share and duration for fatal infections | 37%; 0.8 d | Fixed | Fixed structural assumption |
| $R_0$ | Pre-brake reproduction number | Normal(1.9, 0.5), lower bound 1.2 | 2.63 (2.31–3.08) | Calibrated |
| $k$ | Negative-binomial offspring dispersion | 0.372 | Fixed | Fixed after weak identification in earlier fits |
| $m$ | Probability an offspring leaves the parent’s zone | Beta(2, 12) | 0.19 (0.12–0.29) | Calibrated |
| IFR | Infection fatality ratio | Beta(13.7, 14.3) | 0.46 (0.33–0.59) | Calibrated |
| $\rho_{\text{access}}$ | Spatial access multiplier | Beta(2, 3) | 0.39 (0.10–0.70) | Calibrated |
| $D_{1/2,z}$ | Local visible deaths at half brake strength | Uniform(2, 200) | 113 (44–189) | Calibrated |
| $R_{\text{floor}}$ | Local late-time transmission floor before national scaling | Lognormal(0.04879, 0.30) | 1.08 (0.67–1.76) | Calibrated |
| PCR probability, lab zone | Live-case PCR access with a laboratory | Derived | 0.60 (0.42–0.83) | Derived from calibrated access and collection ratios |
| PCR probability, naive zone | Live-case PCR access without a laboratory or awareness | Derived | 0.29 (0.17–0.45) | Derived from calibrated access and collection ratios |
| Presumptive isolation | Isolation before PCR confirmation | Beta(4, 3) | 0.53 (0.28–0.77) | Calibrated |
| Confirmed isolation | Isolation after PCR confirmation | Ordered uplift over presumptive isolation | 0.71 (0.50–0.88) | Derived from calibrated isolation terms |
| Contact tracing | PCR-confirmed offspring traced and isolated | Beta(5, 2) | 0.69 (0.39–0.90) | Calibrated |
| Baseline postmortem PCR | Status-quo postmortem testing before RDT scale-up | Beta(1, 4) | 0.18 (0.02–0.47) | Calibrated |

## Appendix 2: Posterior Diagnostics

**Figure 8:**
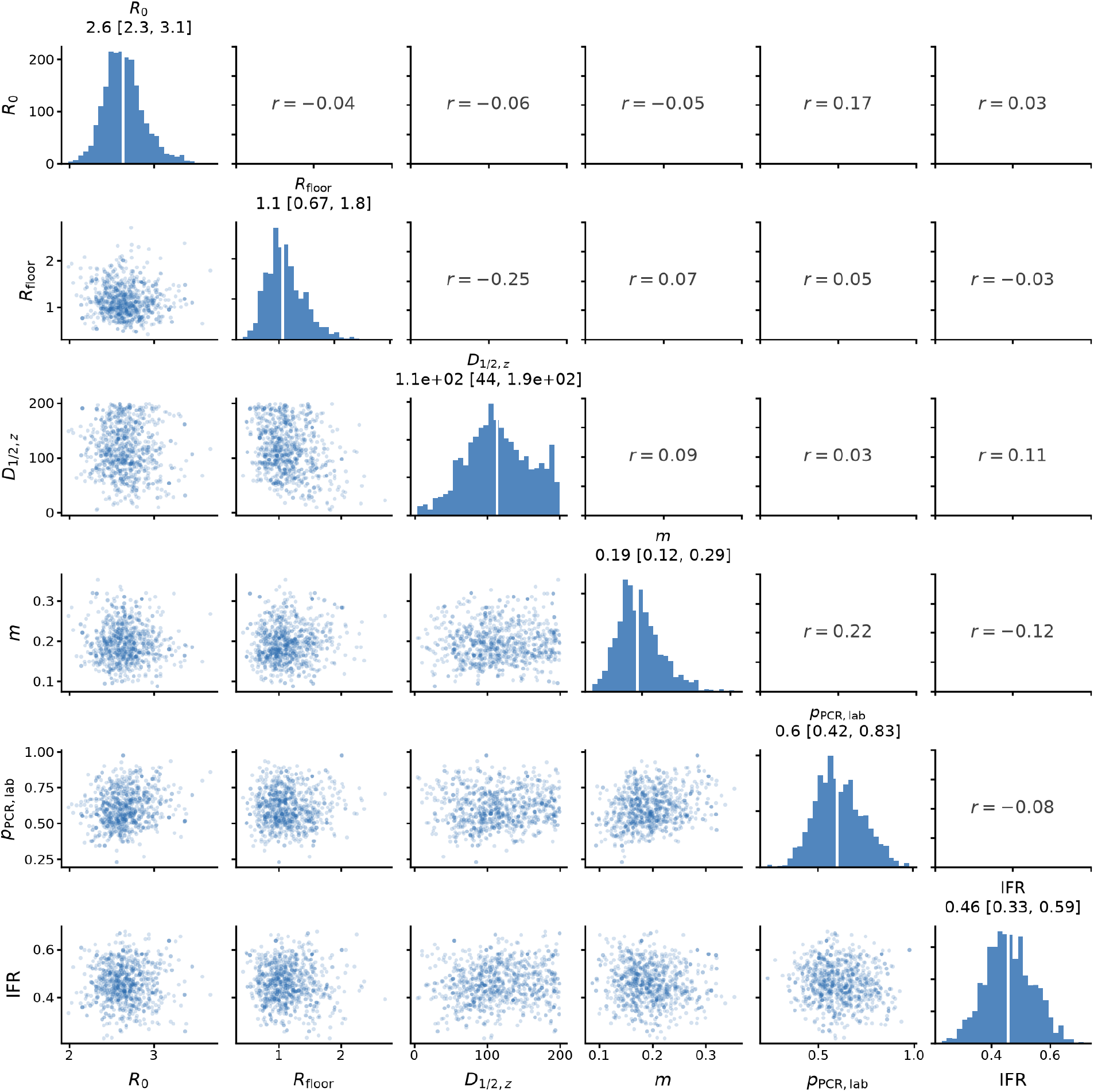
Posterior dependence among selected fitted parameters. Diagonal panels show marginal posterior distributions, lower-triangle panels show paired posterior draws, and uppertriangle panels show Pearson correlations for six parameters that drive the status-quo reproduction number, spatial spread, detection, infection fatality, and local awareness response. Histograms and scatter points are the 2,000 health-zone-only posterior states propagated into the scenario analysis.

**Figure 9:**
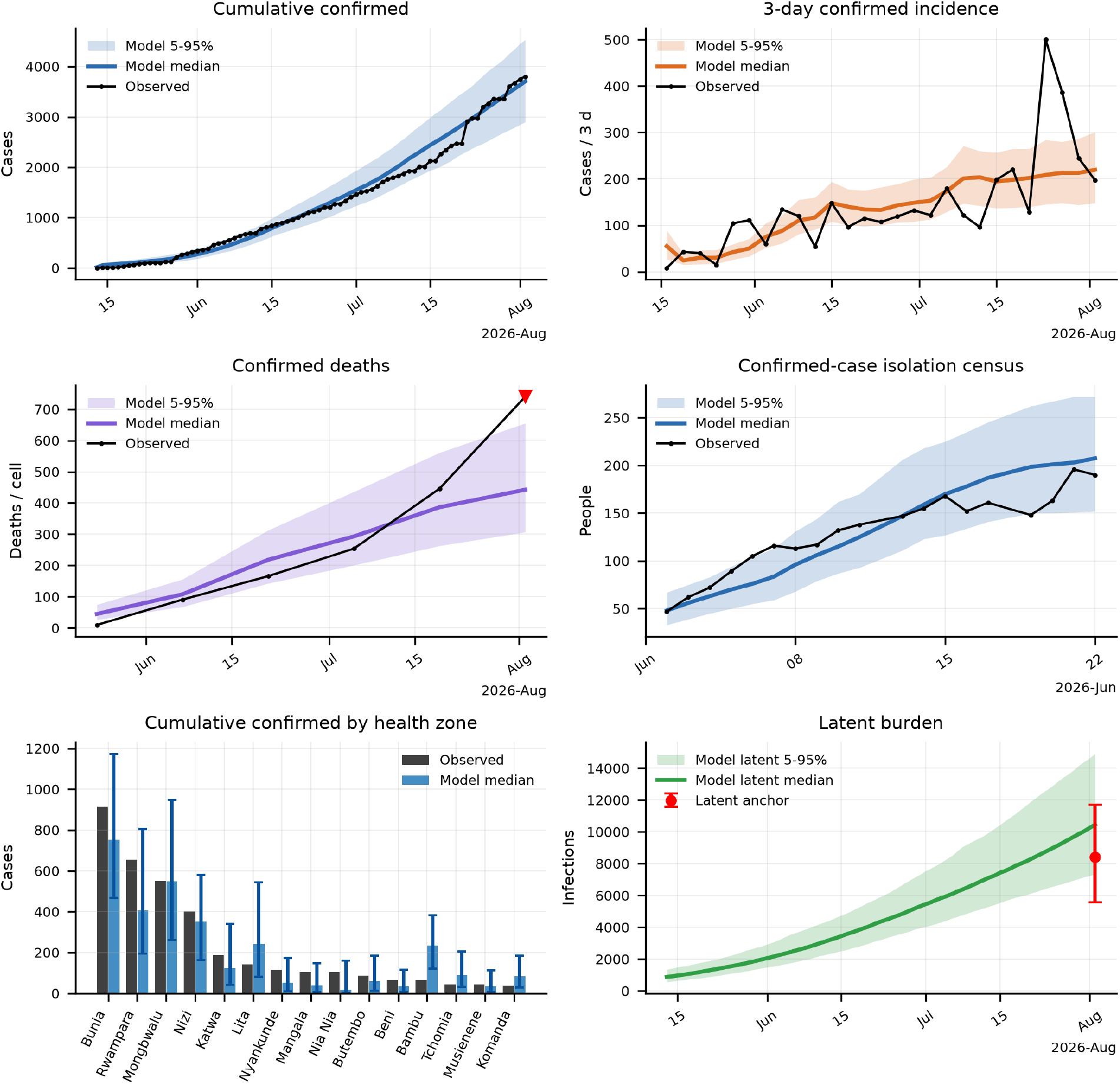
Posterior predictive fit to the calibration data. Bands show 5-95% posterior predictive intervals from the hcalth-zonc-only posterior and black points or lines show the observed calibration targets: cumulative confirmed cases, thrcc-day confirmed incidence, two-wcck confirmed deaths, confirmed-case isolation census, cumulative confirmed cases by high-burden health zone, and the latent-infection anchor.

## Notes

### Competing Interest Statement

The authors have declared no competing interest.

## References

Abbott, S., Sherratt, K., Brand, S., Funk, S., 2026. BVD Outbreak Size: estimating the current size of the 2026 DRC Bundibugyo virus outbreak. URL: https://epiforecasts.io/BVDOutbreakSize/.

Ajelli, M., Mcrlcr, S., Fumanelli, L., Pastore y Piontti, A., Dean, N.E., Longini, I.M., Halloran, M.E., Vespignani, A., 2016. Spatiotemporal dynamics of the Ebola epidemic in Guinea and implications for vaccination and disease elimination: a computational modeling analysis. BMC Medicine 14, 130. doi:10.1186/sl2916-016-0678-3.

Albarino, C.G., Shoemaker, T., Khristova, M.L., Wamala, J.F., Muyembe, J.J., Balinandi, S., Tumusiime, A., Campbell, S., Cannon, D., Gibbons, A., Bergeron, E., Bird, B., Dodd, K., Spiropoulou, C., Erickson, B.R., Guerrero, L., Knust, B., Nichol, S.T., Rollin, P.E., Stroher, U., 2013. Genomic analysis of filoviruses associated with four viral hemorrhagic fever outbreaks in Uganda and the Democratic Republic of the Congo in 2012. Virology 442, 97–100. doi:10.1016/j.virol.2013.04.014.

Andrieu, C., Roberts, G.O., 2009. The pseudo-marginal approach for efficient Monte Carlo computations. The Annals of Statistics 37, 697–725. doi:10.1214/07-A0S574.

Foreman-Mackey, D., Hogg, D.W., Lang, D., Goodman, J., 2013. emcee: The MCMC hammer. Publications of the Astronomical Society of the Pacific 125, 306–312. doi:10.1086/670067.

Huber, C., Watts, A., Thomas-Bachli, A., McIntyre, E., Tuite, A., Khan, K., Cetron, M., Merrill, R.D., 2023. Using spatial and population mobility models to inform outbreak response approaches in the Ebola affected area, Democratic Republic of the Congo, 20182020. Spatial and Spatio-temporal Epidemiology 44, 100558. doi:10.1016/j.sste.2022.100558.

Institut National de Santé Publique, 2026. Rapports de situation. URL: https://insp.cd/category/sitrep/.

Jombart, T., Kada, S., Chakraborty, D., Redding, D.W., Abbate, J., 2026. A stochastic meta-population model of Ebola virus disease transmission for informing public health decisions. Epidemics 55, 100922. doi:10.1016/j.epidem.2026.100922.

Kraemer, M.U.G., Golding, N., Bisanzio, D., Bhatt, S., Pigott, D.M., et al., 2019. Utilizing general human movement models to predict the spread of emerging infectious diseases in resource poor settings. Scientific Reports 9, 5151. doi:10.1038/s41598-019-41192-3.

Kratz, T., Roddy, P., Oloma, A.T., Jeffs, B., Pou Ciruelo, D., de la Rosa, O., Borchert, M., 2015. Ebola virus disease outbreak in Isiro, Democratic Republic of the Congo, 2012: signs and symptoms, management and outcomes. PLOS ONE 10, e0129333. doi:10.1371/journal.pone.0129333.

Kucharski, A.J., Eggo, R.M., Watson, C.H., Camacho, A., Funk, S., Edmunds, W.J., 2016. Effectiveness of ring vaccination as control strategy for Ebola virus disease. Emerging Infectious Diseases 22, 105–108. doi: 10.3201/eid2201.151410.

Leskovec, J., Krause, A., Guestrin, C., Faloutsos, C., VanBriesen, J., Glance, N., 2007. Cost-effective outbreak detection in networks, in: Proceedings of the 13th ACM SIGKDD International Conference on Knowledge Discovery and Data Mining, Association for Computing Machinery. pp. 420–429. doi:10.1145/1281192.1281239.

MacNeil, A., Farnon, E.C., Wamala, J., Okware, S., Cannon, D.L., Reed, Z., Towner, J.S., Tappero, J.W., Lutwama, J., Downing, R., Nichol, S.T., Ksiazek, T.G., Rollin, P.E., 2010. Proportion of deaths and clinical features in Bundibugyo Ebola virus infection, Uganda. Emerging Infectious Diseases 16, 1969–1972. doi:10.3201/eidl612.100627.

McCabe, R., Ebbarnezh, L., Okware, S., Fotsing, R., Koua, E., Mbaka, P., Lofungola, A., van Elsland, S.L., McMenamin, M., Ferguson, N., le Polain de Waroux, O., Cori, A., 2026. Estimation of the size of the outbreak of Ebola disease caused by Bundibugyo virus in the Democratic Republic of the Congo. Technical Report. Imperial College London. URL: https://www.imperial.ac.uk/mrc-global-infectious-disease-analysis/research-themes/preparedness-and-response-to-emerging-threats/report-ebola-18-05-2026/, doi:10.25560/130007.

Raleigh, C., Kishi, R., Linke, A., 2023. Political instability patterns are obscured by conflict dataset scope conditions, sources, and coding choices. Humanities and Social Sciences Communications 10, 74. doi:10.1057/s41599-023-01559-4.

Roddy, P., Howard, N., Van Kerkhove, M.D., Lutwama, J., Wamala, J., Yoti, Z., Colebunders, R., Palma, P.P., Sterk, E., Jeffs, B., Van Herp, M., Borchert, M., 2012. Clinical manifestations and case management of Ebola haemorrhagic fever caused by a newly identified virus strain, Bundibugyo, Uganda, 2007–2008. PLOS ONE 7, e52986. doi:10.1371/journal.pone.0052986.

Towner, J.S., Sealy, T.K., Khristova, M.L., Albarino, C.G., Conlan, S., Reeder, S.A., Quan, P.L., Lipkin, W.I., Downing, R., Tappero, J.W., Okware, S., Lutwama, J., Bakamutumaho, B., Kayiwa, J., Comer, J.A., Rollin, P.E., Ksiazek, T.G., Nichol, S.T., 2008. Newly discovered Ebola virus associated with hemorrhagic fever outbreak in Uganda. PLOS Pathogens 4, e1000212. doi: 10.1371/journal.ppat.1000212.

WHO Ebola Response Team, 2014. Ebola virus disease in West Africa: the first 9 months of the epidemic and forward projections. New England Journal of Medicine 371, 1481–1495. doi:10.1056/NEJMoal411100.

